# Prenatal and Childhood Adverse Events and Child Brain Morphology: A Population-Based Study

**DOI:** 10.1101/2021.02.25.21252442

**Authors:** Andrea P. Cortes Hidalgo, Scott W. Delaney, Stavroula A. Kourtalidi, Alexander Neumann, Runyu Zou, Ryan L. Muetzel, Marian J. Bakermans-Kranenburg, Marinus H. van IJzendoorn, Henning Tiemeier, Tonya White

## Abstract

**Background:** Prenatal and childhood adverse events have been shown to be related to children’s cognitive and psychological development. However, the influence of early-life adversities on child brain morphology is not well understood and most studies are based on small samples and often examine only one adversity. Thus, the goal of our study is to examine the relationship between cumulative exposures to prenatal and childhood adversities and brain morphology in a large population-based study.

**Methods:** Participants included 2,993 children in whom prenatal adversities were reported by mothers at 20-25 weeks of pregnancy and the child’s lifetime exposure to adversities was reported by mothers when the children were 10 years-of-age. The total brain, grey and white matter volumes and the volume of the cerebellum, amygdala and hippocampus were assessed with magnetic resonance imaging when children were 10 years old.

**Results:** In total, 36% of children had mothers who were exposed to at least one adversity during pregnancy and 35% of children were exposed to adversities in childhood. In our study sample, the cumulative number of prenatal adversities was not related to any brain outcome. In contrast, per each additional childhood adverse event, the total brain volume was 0.07 standard deviations smaller (SE = 0.02, p = 0.001), with differences in both grey and white matter volumes. Childhood adversities were not related to the amygdala or hippocampal volumes. Additionally, the link between childhood events and the preadolescent brain was not modified by prenatal events and was not explained by maternal psychopathology.

**Conclusions:** Our results suggest that childhood adversities, but not prenatal adverse events, are associated with smaller global brain volumes in preadolescence. Notably, this is the first large population-based study to prospectively assess the association between the cumulative number of prenatal adversities and the preadolescent brain morphology. The study findings extend the evidence from high-risk samples, providing support for a link between cumulative childhood adverse events and brain morphology in children from the general population.

## Introduction

Adversities, defined as the negative experiences that deviate from the expectable environment, need to be chronic (e.g. parental loss), or sufficiently severe to require a considerable psychobiological adaptation (1). Children whose mothers experienced adversities during pregnancy tend to have more behavioral problems (2) and childhood adversities are associated with poorer intellectual performance (3). Although studies in high-risk samples have addressed the relation between early-life adversity and child brain morphology (1), this association is not well documented in the general population.

Few studies have examined the relation between prenatal adversities and offspring neurodevelopment. As reviewed by Franke, Van den Bergh (4), studies examining head circumference (HC) at birth showed mixed results. For example, prenatal adversities were not related to HC at birth in a population-based sample (N=4,211)(5), whereas a small *positive* association was found in a larger cohort (N=78,017)(6). HC metrics are easily accessible and a proxy for total brain volume. However, they might not capture region-specific differences (4). Only one study assessed prenatal adversities and child brain morphology using magnetic resonance imaging (MRI) and found that girls whose mothers were exposed to an adverse event in pregnancy had *larger* amygdala volumes (N=68) (2). To date, no large population-based study has examined the relation between cumulative prenatal adversities and child brain morphology.

In contrast, there is substantial research on childhood adversities and offspring neurodevelopment, including case-control studies, where adversities are often severe (e.g. institutionalization), and studies in children exposed to a more graded scale of events. Severe adversities have been related to smaller cerebellar (1) and global brain volumes, with differences in multiple brain regions (7). Evidence for differences in the amygdala and hippocampus is mixed, with both larger (8, 9) and smaller volumes (10) reported. Hanson, Nacewicz (10) examined three samples of children exposed to different adversities (physical abuse, neglect, low socioeconomic status (SES)) and found smaller amygdala in relation to all adversities.

Studies in children exposed to more common adversities have reported differences in the cerebellum, cortex and limbic structures. Cumulative childhood adversities were related to smaller cerebellar volumes in a sample of 58 adolescents (11) and to smaller prefrontal cortex, amygdala and hippocampal volumes in a study oversampled for child depression (12, 13). Importantly, the adversity definition in the latter study included parental psychopathology. Although having a parent with psychopathology may represent an adversity, shared genetic factors may underlie the association (4) and parental psychopathology may additionally interact with the adversities’ effect (14).

Previous studies (7, 10) described similar findings in relation to different adversities, which could imply a low specificity across adversity types. Evidence also suggests a cumulative relation between childhood adversities and numerous outcomes, including health-risk behaviors and psychiatric disorders (15). Thus, we assessed the cumulative effect of early-life adversities on brain morphology. Compared to examining single adversities, the cumulative adversity assessment (15) offers a more naturalistic view of the adversity exposure, because adverse events are often related and tend to co-occur (16).

Notably, a randomized-controlled trial in institutionalized children demonstrated that cognitive outcomes improved when children were placed into foster care, especially if this placement occurred at younger ages (3). Bick and Nelson (7) additionally described smaller gray matter volumes in children ever-institutionalized compared to those never-institutionalized. Thus, child neurodevelopment can improve, within the available biological reserve, after adversity ceases (17). This has two implications for our study. First, the timing of adversity exposure may influence the association with brain morphology. Children with no childhood adversities, but whose mothers experienced adversities during pregnancy may show differences due to the pronounced neurodevelopment that occurs during prenatal life (17). Children with adversities in both the prenatal and childhood periods may have the largest brain differences.

Thus, we examined adversities in both periods in relation to child brain morphology. Second, when adversity occurs only prenatally, the brain development could “catch-up”, approaching the typical growth curve (17). To examine this neuroplasticity, we included fetal HC measures in sensitivity analyses.

In this population-based study, we examined the relationship between cumulative prenatal and childhood adversities and the preadolescent brain morphology. We hypothesized a greater number of adversities would be related to smaller global brain, amygdala and hippocampal volumes. We additionally hypothesized a stronger association between childhood adversities and brain morphology in children whose mothers were exposed to prenatal adversities.

## Materials and Methods

### Participants

This study is part of the Generation R Study, a population-based prenatal birth cohort in Rotterdam, the Netherlands (18). In total, 9,778 pregnant mothers with a delivery date from April 2002 to January 2006 were enrolled, and information was collected from children and parents by questionnaires, interviews and research visits. Study protocols for each wave of data collection were approved by the Medical Ethical Committee of the Erasmus Medical Center and all parents gave written informed consent.

T_1_-weighted MRI scans were acquired in 3,966 9-to-11-year-old children (19), of which 3,186 had good image quality data. Among these children, 3,146 had complete information on prenatal and/or childhood adversities. We randomly excluded one sibling (N=153) to avoid non-independent data. In total, 2,993 children were included in analyses (2,242 in prenatal adversities analyses and 2,923 in childhood adversities analyses; Figure S1).

### Measures

#### Prenatal adversities

Adverse events occurring prenatally and shortly before pregnancy were assessed with a Dutch-adapted version of the Social Readjustment Rating Scale (SRRS)(20). At 20-25 weeks of pregnancy, mothers reported the occurrence of ten stressful events in the preceding 12 months (e.g. serious illnesses of family members, partner’s death)(21). An additional question on the occurrence of robbery, theft, physical abuse or rape was also included, given the relevance of these adverse experiences. Moving to a new home, originally assessed by the SRRS, was excluded as it could also reflect a positive situation. A ‘*prenatal adversities score*’ was computed as the cumulative number of occurrences of ten adverse events (Table S1).

#### Childhood adversities

Occurrence of stressful life events from birth to age 10 years was reported by mothers during an interview when children were 10 years old (22). This instrument was based on the TRAILS study questionnaires (23) and the Life Events and Difficulty Schedule (24), and comprised twenty-four events of varying severity (e.g. high school workload, parental conflicts). To better measure *severe* adversities in this population-based sample, specific adverse events were selected using as reference the ACEs studies (e.g. Felitti, Anda (15)). A ‘*childhood adversities score*’ was computed as the cumulative occurrence of these adversities (Table S2).

### Brain Imaging

Brain MRI data were obtained in 9-11-year-old children using a 3 Tesla GE 750w Discovery platform (General Electric, Milwaukee, WI)(19). T_1_-weighted images were collected with a receive-only 8-channel head coil and an inversion recovery fast spoil gradient recalled sequence (TR=8.77ms, TE=3.4ms, TI=600ms, Flip angle=10°, Field of view=220×220, Acquisition matrix=220×220, Slice thickness=1mm, Number of slices=230, ARC acceleration factor=2).

We processed and conducted the segmentation and reconstruction of the neuroimaging data with the FreeSurfer image analysis suite (v.6.0)(25). Reconstructed images were inspected for quality and poor quality reconstructions were excluded from further analyses (26). The total brain volume, the cortical grey and cerebral white matter volumes, the cerebellar volume, and the amygdala and hippocampal volumes were included in analyses.

### Ultrasound measures

Fetal ultrasound measures were collected at three time-points during pregnancy (27). Trained sonographers established the gestational age based on the first ultrasound assessment and measured fetal HC using standardized techniques (28). The HC measures collected during the third trimester of pregnancy were included in sensitivity analyses.

### Covariates

We included as covariates child sex and age at the MRI scan, total intracranial volume, maternal ethnicity, highest household education and maternal prenatal alcohol use and smoking. Child sex was collected from birth records. Maternal ethnicity was defined based on her parents’ birth country and was self-reported during pregnancy. The highest household education, and prenatal alcohol consumption and smoking were reported through questionnaires during pregnancy (See Supplemental Information).

Maternal psychopathology in pregnancy was assessed with the Brief Symptom Inventory, a validated and widely-used questionnaire (29). We used the global severity index score, a measure of the global severity of psychopathology, in additional analyses.

### Statistical Analyses

We examined the associations of prenatal and childhood adversities with the brain outcomes using multiple linear regression. We first fitted a minimally adjusted model controlling for child sex and age at MRI scan, total intracranial volume (in amygdala and hippocampus analyses) and maternal ethnicity. To account for confounding by SES, we additionally adjusted for the highest household education in a second model. Finally, we also controlled for prenatal alcohol use and smoking in a third, fully-adjusted model, since these factors could be also considered part of the pathway between prenatal adversities and brain morphology.

We subsequently examined the interaction between prenatal and childhood adversities in relation to brain morphology. Additionally, for descriptive purposes, we assessed the relation between a categorical adversity measure and the brain outcomes, using four groups: children with one or more prenatal adversities *only* (N=460), children with one or more childhood adversities *only* (N=433), children with adversities in both periods (N=321), and children with no adversities (N=958).

Several sensitivity analyses were performed. We first examined whether child sex modified the associations between adversity and brain morphology. Second, we analyzed the associations of adversity and brain morphology in a more homogeneous group, children whose mothers had a Dutch national origin. Third, we explored whether associations between adversity and brain morphology were explained by maternal psychopathology, and we examined the interaction between maternal psychopathology and adversity in relation to child brain morphology. Finally, to explore the role of brain plasticity (17), we assessed whether prenatal adversities were associated with HC at the last pregnancy trimester, as HC is a proxy for an early measure of total brain volume (analyses adjusted for gestational age at ultrasound).

Analyses were performed in R v.3.6.1 (30). Outcomes were standardized. Multiple imputation of missing values (maximum missingness: maternal psychopathology=23.4%) was performed (“mice” package (31)), and results were pooled across 25 imputed datasets. We found no signs of violation of the regression assumptions (i.e. independence, normal distribution, homoscedasticity). Adjustment for multiple testing was performed using the FDR approach (32) in the analyses with prenatal adversities, childhood adversities and the interaction between prenatal and childhood adversities (15 tests, including all brain outcomes, except for total brain volume).

### Non-response analyses

Children included in the analyses of prenatal adversities and brain morphology (N=2,242) were compared to children with data on prenatal adversities but no neuroimaging data available (N=3,552). Continuous variables were compared with the Mann-Whitney U test and categorical variables with chi-squared tests. Mothers of children without imaging data were more often exposed to prenatal adversities (one or more events: 40.7%) than those of children in analyses (one or more events: 36.1%) and were less often highly educated (22.1% vs 30.5%). Additionally, mothers of children without imaging data were less often from Dutch origins (No imaging data group: 50.6%; Study sample: 61.1%) and had more psychiatric symptoms (median (IQR)=0.19 (0.1, 0.4)) than those in analyses (median (IQR)=0.15 (0.1, 0.3)).

## Results

In total, 36% of children had mothers who were exposed to at least one prenatal adversity and 35% of children were exposed to adversities during childhood (Table 1). The most commonly reported prenatal event was a substantial financial downturn (14.5%), followed by a serious illness of a family member (11.6%)(Table S1). In childhood, parental separation or divorce was the most prevalent event (21.45%)(Table S2). Children with mothers exposed to prenatal adversities were more likely to experience adversities during childhood (41%) compared to those without prenatal adversities (31%).

**Table 1.** Baseline characteristics

|  | mean (SD) or<br>%* | N |
| --- | --- | --- |
| **Adversity measures** |  |  |
| Prenatal adversities (10 items), % (N=2242) |  |  |
| 0 | 63.9 | 1432 |
| 1 | 20.6 | 461 |
| 2 | 10.5 | 236 |
| 3 | 3.8 | 85 |
| 4 or more | 1.2 | 28 |
| Childhood adversities (4 items), % (N=2923) |  |  |
| 0 | 64.9 | 1897 |
| 1 | 27.2 | 795 |
| 2 | 6.3 | 185 |
| 3 | 1.4 | 41 |
| 4 | 0.2 | 5 |
| **Child characteristics** |  |  |
| Sex, % girls | 50.8 | 1521 |
| Age at MRI scan, years | 10.1 (0.6) | 2993 |
| **Parental Characteristics** |  |  |
| Maternal ethnicity, % |  | 2993 |
| Dutch | 57.6 | 1725 |
| Non-Western | 30.3 | 906 |
| Other Western | 12.1 | 362 |
| Highest household education, % |  | 2993 |
| Low education | 41.0 | 1227 |
| Medium education | 22.4 | 670 |
| High education | 36.6 | 1096 |
| Maternal prenatal alcohol use, % never during pregnancy | 41.0 | 1226 |
| Maternal prenatal smoking, % never during pregnancy | 76.9 | 2303 |
| Maternal Psychiatric Symptoms, median (Q1,Q3) | 0.15 (0.06, 0.32) | 2993 |

Characteristics of the sample with information for **prenatal AND/OR childhood adversities** and brain structural MRI data (N=2993). \*Otherwise indicated. Based on imputed datasets.

The cumulative number of prenatal adverse events was not related to any brain outcome (Table 2). In contrast, a consistent association was found between childhood adversities and all global brain metrics (total brain, cortical grey and white matter volumes and total cerebellar volumes). Children had, on average, a 0.11 standard-deviation smaller total brain volume (SE=0.02, p<0.001) per each additional childhood adverse event, adjusting for child sex, age at the MRI scan, and maternal ethnicity. The associations between childhood adversities and the total brain, cortical grey and white matter volumes remained after adjustment for parental education, and prenatal alcohol use and smoking (Total brain volume: B=-0.07, SE=0.02, p=0.001). Childhood adversities were not related to the amygdala and hippocampus (Table 2). After adjustment for multiple testing, the associations between childhood adversities and the cortical grey (p-adjusted=0.02), and cerebral white matter volumes (p-adjusted=0.02) remained.

**Table 2.** Associations between cumulative prenatal and childhood adversities and child brain morphology

|  | Model 1 |  |  | Model 2 |  |  | Model 3 |  |  |
| --- | --- | --- | --- | --- | --- | --- | --- | --- | --- |
|  | B | SE | P | B | SE | P | B | SE | P |
| ***Prenatal adversities*** |  |  |  |  |  |  |  |  |  |
| *Global brain metrics* |  |  |  |  |  |  |  |  |  |
| Total brain volume | -0.03 | 0.02 | 0.14 | -0.02 | 0.02 | 0.39 | -0.01 | 0.02 | 0.52 |
| Cortical grey matter volume | -0.03 | 0.02 | 0.20 | -0.01 | 0.02 | 0.57 | -0.01 | 0.02 | 0.71 |
| Cerebral white matter volume | -0.02 | 0.02 | 0.23 | -0.02 | 0.02 | 0.41 | -0.01 | 0.02 | 0.56 |
| Total cerebellar volume | -0.03 | 0.02 | 0.10 | -0.03 | 0.02 | 0.20 | -0.02 | 0.02 | 0.26 |
| *Subcortical brain metrics* |  |  |  |  |  |  |  |  |  |
| Amygdala, mean volume | 0.02 | 0.02 | 0.40 | 0.01 | 0.02 | 0.41 | 0.01 | 0.02 | 0.52 |
| Hippocampus, mean volume | 0.01 | 0.02 | 0.42 | 0.01 | 0.02 | 0.42 | 0.01 | 0.02 | 0.50 |
| ***Childhood adversities*** |  |  |  |  |  |  |  |  |  |
| *Global brain metrics* |  |  |  |  |  |  |  |  |  |
| Total brain volume | -0.11 | 0.02 | <0.001 | -0.08 | 0.02 | <0.001 | -0.07 | 0.02 | 0.001 |
| Cortical grey matter volume | -0.11 | 0.02 | <0.001 | -0.08 | 0.02 | 0.001 | -0.07 | 0.02 | 0.003* |
| Cerebral white matter volume | -0.10 | 0.02 | <0.001 | -0.08 | 0.02 | 0.001 | -0.07 | 0.02 | 0.002* |
| Total cerebellar volume | -0.07 | 0.02 | 0.003 | -0.05 | 0.02 | 0.03 | -0.05 | 0.02 | 0.06 |
| *Subcortical brain metrics* |  |  |  |  |  |  |  |  |  |
| Amygdala, mean volume | 0 | 0.02 | 0.90 | 0 | 0.02 | 0.87 | -0.01 | 0.02 | 0.70 |
| Hippocampus, mean volume | -0.01 | 0.02 | 0.58 | -0.01 | 0.02 | 0.63 | -0.01 | 0.02 | 0.59 |

Model 1 adjusted for child age at MRI scan, child sex, total intracranial volume (in subcortical metrics), maternal ethnicity. Model 2 additionally adjusted for the highest household education. Model 3 additionally adjusted for maternal prenatal alcohol use and smoking. All outcomes are standardized. N=2242 in prenatal adversities analyses, N=2923 in childhood adversities analyses. \*These p-values survived correction for multiple testing.

No interaction was observed between prenatal and childhood adversities in relation to child brain morphology (Table 3). Also, when using the categorical adversity measure, the exposure to *only* prenatal adversities was not related to the total brain volume, whereas the specific exposure to childhood adversities was associated with a 0.10 standard-deviation smaller total brain volume (p=0.04). Additionally, children with adversities in both periods had a 0.10 standard-deviation smaller total brain volume than those non-exposed (p=0.06). Altogether, our results suggest that only childhood events are related to brain morphology and that this association is independent of the occurrence of prenatal adversities (Figure 1).

**Table 3.** Interaction between prenatal adversities and adversities in childhood in relation to brain morphology

|  | Main effect:<br>Prenatal<br>adversities |  |  | Main effect:<br>Adversities in<br>childhood |  |  | Interaction Effect |  |  |
| --- | --- | --- | --- | --- | --- | --- | --- | --- | --- |
|  | B | SE | P | B | SE | P | B | SE | P |
| *Global metrics* |  |  |  |  |  |  |  |  |  |
| Total brain volume | -0.02 | 0.02 | 0.33 | -0.10 | 0.03 | 0.002 | 0.04 | 0.03 | 0.15 |
| Cortical grey matter volume | -0.02 | 0.02 | 0.33 | -0.10 | 0.03 | 0.001 | 0.04 | 0.03 | 0.08 |
| Cerebral white matter volume | -0.02 | 0.03 | 0.55 | -0.09 | 0.03 | 0.01 | 0.02 | 0.03 | 0.40 |
| Total cerebellar volume | -0.03 | 0.03 | 0.22 | -0.06 | 0.03 | 0.09 | 0.03 | 0.03 | 0.35 |
| *Subcortical metrics* |  |  |  |  |  |  |  |  |  |
| Amygdala, mean volume | 0 | 0.02 | 0.96 | -0.02 | 0.03 | 0.43 | 0.03 | 0.02 | 0.24 |
| Hippocampus, mean volume | 0.01 | 0.02 | 0.56 | 0.01 | 0.03 | 0.69 | 0 | 0.02 | 0.94 |

Model adjusted for child age at MRI scan, child sex, total intracranial volume (in subcortical metrics), maternal ethnicity, the highest education in the household, maternal prenatal alcohol use and maternal prenatal smoking. All brain outcomes were standardized. Adversities measures represent the cumulative number of events. N=2172

**Figure 1.**
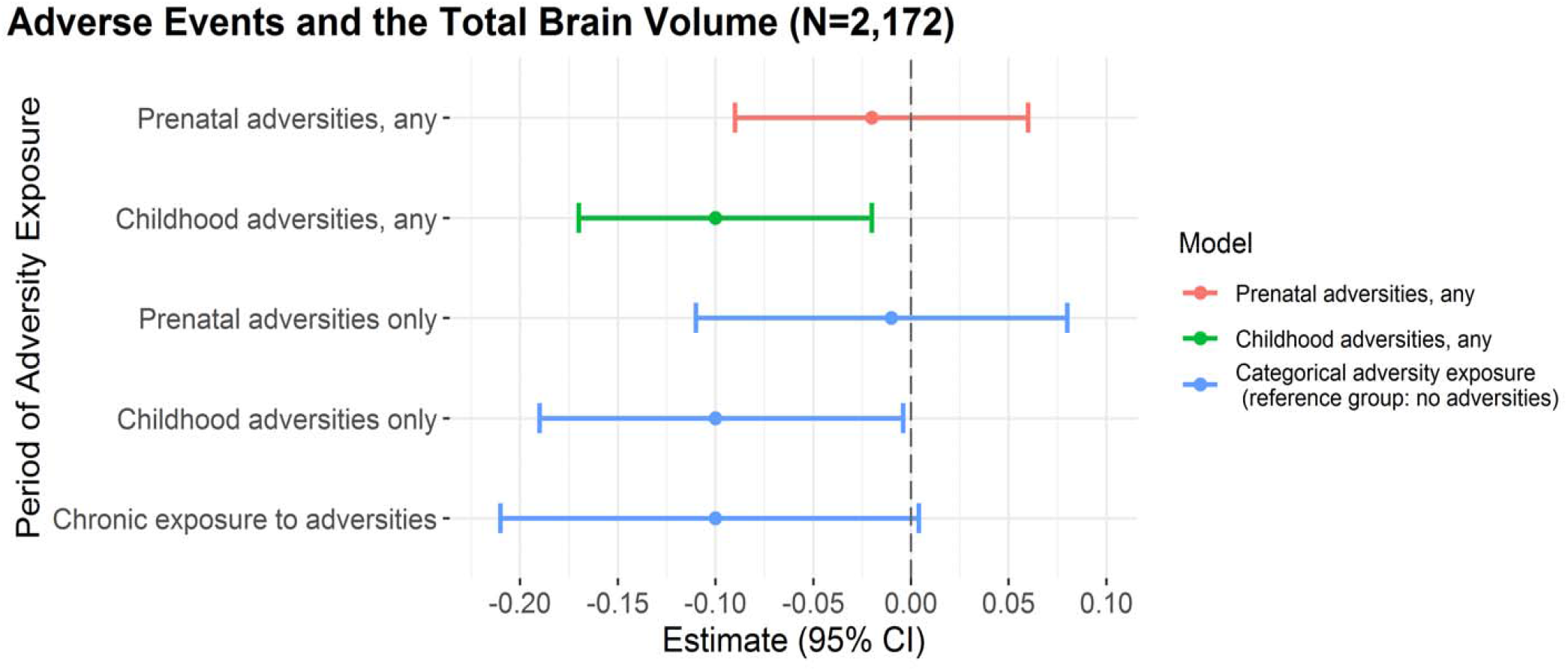
Associations between prenatal and childhood adversities with the total brain volume.

We further examined the specificity of the association between childhood adversities and brain morphology. No interaction was found between child sex and childhood adversities for any brain outcome. When including only children with Dutch mothers, childhood adversities were related to the total brain, grey and white matter, and cerebellar volumes (Table S3). Also, the associations between childhood adversities and brain morphology were not explained nor modified by maternal prenatal psychopathology (Table S4). Finally, the cumulative number of prenatal adversities was not related to variations in the fetal HC (B=0.00, SE=0.02, p=0.82; N=2,168).

## Discussion

In this population-based study, childhood adversities, but not prenatal adverse events experienced by the mother, were related to global brain volume differences at age 10 years. Our study provides two novel contributions to the literature. This is the first study to examine the association between cumulative prenatal adversities and brain structure in children from the general population. Contrary to our hypothesis, we found no relationship between cumulative prenatal adversities and preadolescent brain morphology using a large population-based sample, an assessment of prenatal adversities while mothers were pregnant and neuroimaging data. Second, cumulative childhood adversities were related to smaller total brain volumes and differences were observed across grey and white matter volumes. These findings are consistent with research in some small high-risk samples, supporting a relation between cumulative childhood adversities and child neurodevelopment.

The absence of associations between prenatal adversities and child brain morphology is surprising, as the brain undergoes dramatic developmental changes during pregnancy (17). Our study may have lacked sufficient power to observe subtle effects. However, we assessed a considerably larger sample than previous studies (2). The brain can adapt in response to environmental effects (7), which raises the question of whether brain plasticity could have obscured an association between prenatal adversities and brain morphology. Given a rich and positive childhood environment, the brain development of children whose mothers experienced stress in pregnancy could catch-up and return to the normative trajectory (17). If this were the case, prenatal adversities would be related to brain differences earlier in life. However, we found no association between prenatal events and HC in the last pregnancy trimester, arguing against the plasticity hypothesis (17) (see also a study from this cohort examining family dysfunction and fetal HC (27)). It is also possible that the adversity type and severity influence the relation with brain morphology. Whereas Jones, Dufoix (2) found a relation between the gestational exposure to a natural disaster and amygdala volumes, the cumulative exposure to a range of more normative adverse events was not associated with the global brain volume nor the amygdala and hippocampus in our study.

Numerous studies have examined *childhood* adversity and brain morphology, but results are difficult to compare due to differences in the events assessed, the age of occurrence of adversities and the age at the MRI assessment (7). Overall, research suggests that children exposed to early-life adversity have smaller total brain, grey and white matter, and cerebellar volumes (7). Consistently, we observed that childhood adversity was related to smaller total brain volumes, and this finding was robust to the adjustment for confounders. Analyses with the grey and white matter volumes further supported this association. Additionally, maternal psychopathology did not explain nor modify the relation between childhood adversity and these brain outcomes.

Contrary to what we expected, childhood adversities were not related to the limbic volumes. The amygdala and hippocampus are of particular interest because they have a high density of cortisol receptors and cortisol influences the neuronal development (4). Interestingly, both larger and smaller amygdala and hippocampal volumes have been reported (8–10). In addition to the methodological differences across studies, various hypotheses could underlie these mixed findings. The volumetric growth of the amygdala and hippocampus peaks at around age 10 years (33), thus different findings could be expected between studies assessing brain morphology during childhood, preadolescence, and at later ages. The adversity severity may also influence the results, and the impact of early adversity in some structures may only become apparent later in development (7). Further, the amygdala (34) and hippocampus (35) show continued neurogenesis after fetal life, suggesting that these regions could undergo plastic changes in response to adversity and other environmental factors.

Our adversity measures were selected with a focus on *concrete* environmental events, that could generate stress in the pregnant mother or the child and require a substantial psychobiological adaptation (1). The cumulative prenatal adversity measure was based on a major life events inventory (20), similar to those included in other population-based studies (36). Similarly, our childhood adversity measure included events assessed by key childhood adversities studies (15, 37), previously shown to be associated with greater child psychopathology (22). Different items were used in the prenatal and childhood adversity measures, to focus on *maternal* stressful events in the prenatal measure, and on *childhood* adverse events in the latter measure. Consistent with previous studies (36), the cumulative exposure to prenatal adversities was related to the number of childhood adversities.

Our study has some limitations. First, we did not account for the age of occurrence of childhood adversities. Although events at specific ages could have different effects in brain morphology, it is difficult to determine the exact period of occurrence of adversities that are often chronic and variable (2). Second, mothers reported childhood adversities at age 10 years and thus these reports could be affected by recall bias. Nonetheless, other methods to collect information on childhood adversity in the general population, such as adolescent reports, are limited by the accuracy in reporting early-life events (8). Third, mothers of children without imaging data were more often exposed to prenatal adversities and were less often highly educated than mothers in our study. Finally, by including events that occurred before pregnancy, we could have miss-classified some women who were not experiencing prenatal stress as exposed. However, cumulative preconception adversities have also been shown to predict poor offspring outcomes (38).

## Conclusion

In conclusion, we found that the number of adversities experienced by the mother during pregnancy was not related to brain morphological differences in children from the general population. Childhood adversities consistently predicted smaller brain volumes, with alterations in both grey and white matter volumes. The association between childhood adversities and the global brain volume was not modified by maternal psychopathology, nor by the number of prenatal adversities. Our results support a cumulative association between childhood adversities and brain morphology, previously described in small high-risk samples. If replicated in large samples with repeated MRI and adversity assessments, priority should be given to intervention studies that determine whether providing additional support to children following periods of adversities will prevent the emergence of brain differences.

## Data Availability

The datasets analyzed in this study are currently not publicly available due to legal and ethical restraints due to the General Data Protection Regulations (GDPR). However the consent has been altered for the current wave of data collection which will provide the participants the option to determine the extent that they want their data shared. Via data transfer agreements, the data can be made available upon request. Interested researchers can direct their requests to Vincent Jaddoe.

## Ethics Statement

All study protocols and the measurements assessed in each wave of data collection were approved by the Medical Ethical Committee of the Erasmus MC, University Medical Center Rotterdam.

## Acknowledgements

This work was supported by the Netherlands Organization for Health Research and Development (ZonMw TOP 91211021), Spinoza prize, the National Institutes of Health (F31HD096820), the Harry Frank Guggenheim Foundation, the Dutch Ministry of Education, Culture, and Science (NWO 024.001.003 and 016.VICI.170.200), the Canadian Institutes of Health Research, the European Research Council (ERC-AdG.669249), the China Scholarship Council (201606100056), the Sophia Foundation (S18-20), NWO Physical Sciences Division and SURFsara. The Generation R Study is supported by Erasmus MC, the Erasmus University Rotterdam, ZonMw, NWO, and the Ministry of Health, Welfare and Sport.

