## Supplementary Methods for "Prenatal and Childhood Adverse Events and Child Brain Morphology: A Population-Based Study"

**Covariates**

Maternal ethnicity was categorized as Dutch, non-Dutch Western and non-Western (1). Alcohol consumption during pregnancy included four categories: “never during pregnancy”, “until pregnancy was known”, “continued drinking occasionally in pregnancy”, “continued drinking frequently in pregnancy”. Maternal prenatal smoking was categorized into: “never during pregnancy”, “until pregnancy was known” and “continued in pregnancy”. Information on maternal and paternal education was collected by self-report during pregnancy and was classified following the Dutch standard classification of education (2). The highest education in the household was included in analyses.

**Additional References**

1.	Statistics Netherlands. Statistical Yearbook of the Netherlands 2004. 2004.

2.	Statistics Netherlands. Standaard Onderwijsindeling 2003 Statistics Netherlands (Centraal Bureau voor de Statistiek): Voorburg/Heerlen, the Netherlands; 2005.
