## Supplementary Figures and Tables for "Prenatal and Childhood Adverse Events and Child Brain Morphology: A Population-Based Study"

**Supplementary Figure 1**. Flowchart of sample selection

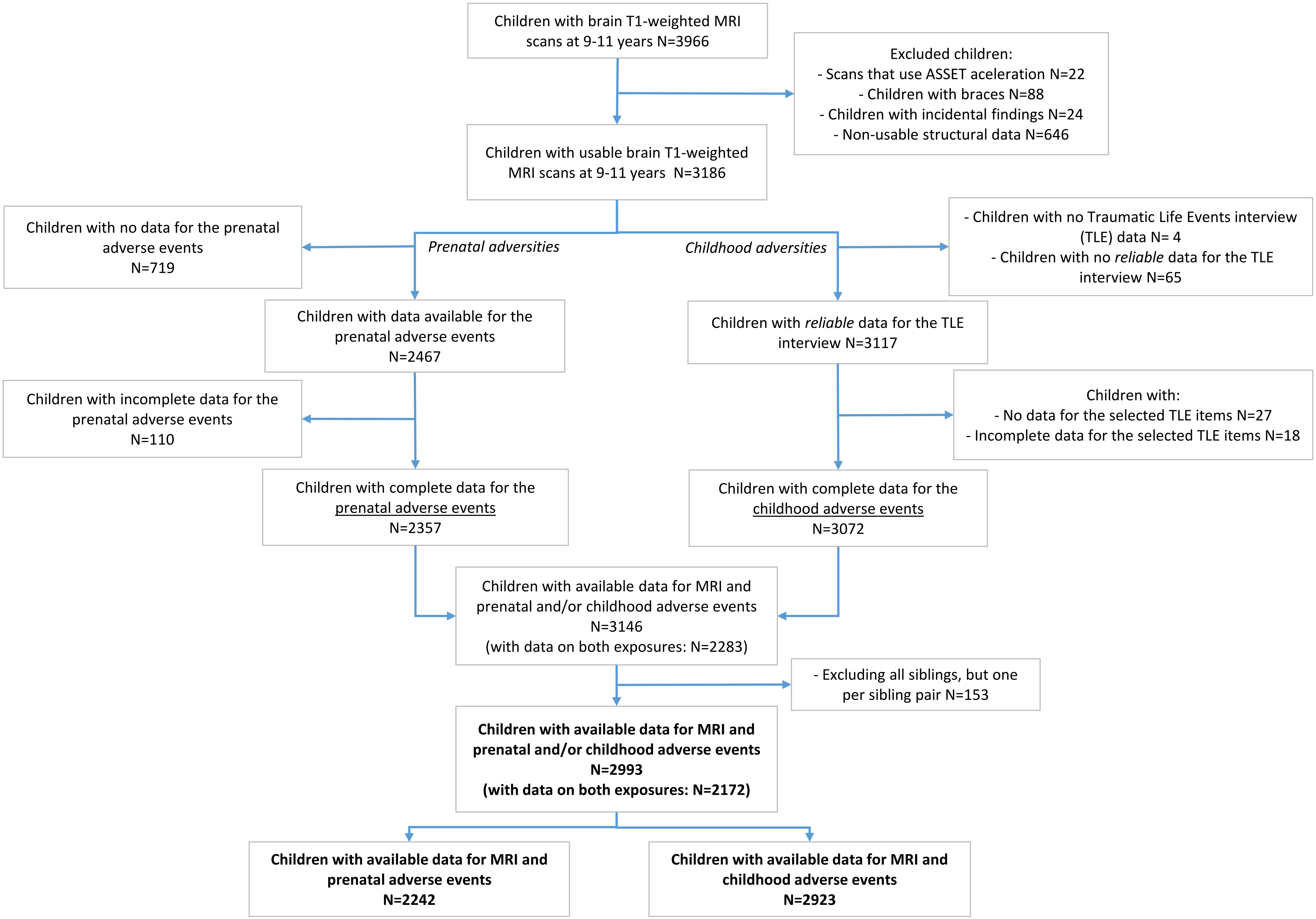

**Supplementary Table 1.** Prevalence of prenatal adverse events

| Event | Prevalence, % | N exposed |
|---|---|---|
| Have you been a victim of robbery, theft, physical abuse or rape? | 3.88 | 87 |
| Have you suffered a substantial downturn in your financial situation? | 14.5 | 325 |
| Have you become unemployed? | 8.97 | 201 |
| Has your partner or other member of your family become unemployed? | 6.51 | 146 |
| Has one or more of your children been seriously ill? | 1.52 | 34 |
| Has your partner, or other family member, or one of your parents (in-law) been seriously ill? | 11.6 | 260 |
| Has one of your children died? | 0.71 | 16 |
| Has your partner died? | 0.04 | 1 |
| Has your father or mother (in-law), a brother or sister, or good friend died? | 7.09 | 159 |
| Have you had a divorce or broken off the relationship with your partner? | 3.57 | 80 |
| ***Any category reported*** | 36.13 | 810 |

N=2242

**Supplementary Table 2.** Prevalence of childhood adverse events

| Category of childhood exposure | Prevalence per category, % | N exposed |
|---|---|---|
| **Psychological abuse** | | |
| Has anyone almost used physical violence against your child? So that it did not actually happen, but your child was scared. | 11.53 | 337 |
| **Physical abuse** | | |
| Has anyone ever used physical violence against your child? For example, beating him/her up. | 6.77 | 198 |
| **Sexual abuse** | 4.41 | 129 |
| Has anyone made sexual comments or movements towards your child?* | 3.42 | 100 |
| Did your child experience inappropriate sexual behavior?* | 1.61 | 47 |
| **Parental loss** | 22.03 | 644 |
| Is your child's father / mother or other caregiver still alive? (reversed)* | 0.89 | 26 |
| Are you and your partner divorced or separated?* | 21.45 | 627 |
| ***Any category reported*** | 35.1 | 1026 |

N=2923. * Sexual abuse and parental loss categories include two items.

**Supplementary Table 3**. Association between childhood adversities and brain morphology in children with Dutch mothers

| | B | SE | P |
|---|---|---|---|
| **Outcome** | | | |
| *Global metrics* | | | |
| Total brain volume | -0.09 | 0.03 | 0.004 |
| Cortical grey matter volume | -0.08 | 0.03 | 0.02 |
| Cerebral white matter volume | -0.09 | 0.03 | 0.01 |
| Total cerebellar volume | -0.08 | 0.03 | 0.02 |
| *Subcortical metrics* | | | |
| Amygdala, mean volume | 0 | 0.03 | 0.94 |
| Hippocampus, mean volume | -0.03 | 0.03 | 0.37 |

Analyses performed in children with Dutch mothers. Model adjusted for child age at MRI scan, child sex, total intracranial volume (in subcortical metrics), the highest education in the household, maternal prenatal alcohol use and maternal prenatal smoking. All outcomes are standardized. N=1669.

**Supplementary Table 4**. Interaction between maternal psychopathology and childhood adversities in relation to child brain morphology

| | Interaction effect | | |
|---|---|---|---|
| | **B** | **SE** | **P** |
| **Outcome** | | | |
| *Global metrics* | | | |
| Total brain volume | 0.08 | 0.06 | 0.19 |
| Cortical grey matter volume | 0.08 | 0.06 | 0.23 |
| Cerebral white matter volume | 0.09 | 0.07 | 0.19 |
| Total cerebellar volume | 0.02 | 0.06 | 0.78 |
| *Subcortical metrics* | | | |
| Amygdala, mean volume | 0.01 | 0.06 | 0.91 |
| Hippocampus, mean volume | 0.02 | 0.06 | 0.74 |

Model adjusted for child age at MRI scan, child sex, total intracranial volume (in subcortical metrics), maternal ethnicity, the highest education in the household, maternal prenatal alcohol use, maternal prenatal smoking, maternal psychiatric symptoms and the interaction term of maternal psychiatric symptoms with childhood adversities. All outcomes are standardized. N=2923
